# Risk of Suicide Attempts and Self-Directed Violence after SARS-CoV-2 Infection: Outcomes from an Emulated Trial of a Nationwide Observational Matched Cohort of US Veterans

**DOI:** 10.1101/2022.12.23.22283902

**Authors:** Denise M. Hynes, Meike Niederhausen, Jason Chen, Troy A. Shahoumian, Mazhgan Rowneki, Alex Hickok, Megan Shepherd-Banigan, Eric J. Hawkins, Jennifer Naylor, Alan Teo, Diana J. Govier, Kristin Berry, Holly McCready, Thomas F. Osborne, Edwin Wong, Paul L. Hebert, Valerie A. Smith, C. Barrett Bowling, Edward J. Boyko, George N. Ioannou, Theodore J. Iwashyna, Matthew L. Maciejewski, Ann M. O’Hare, Elizabeth M. Viglianti, Amy S-B. Bohnert, the VA HSR&D COVID Observational Research Collaboratory (CORC)

## Abstract

**Importance:** The negative health-related effects of SARS-CoV-2 infection may include increased risk for self-directed violence.

**Objective:** To assess suicide attempts and other self-directed violence risk among US Veterans with a positive polymerase chain reaction (PCR) test for SARS-CoV-2 infection compared to matched uninfected Veterans.

**Design, Setting, and Participants:** Using a target trial emulation design supported by comprehensive electronic health records from the US Veterans Health Administration, Veterans who had a positive PCR test between March 1, 2020 and March 31, 2021 were matched with non-infected comparators. Monthly matching was anchored to first positive PCR test for each patient. Groups were followed for one-year thereafter.

**Exposure:** Positive SARS-CoV-2 PCR.

**Main Outcomes and Measures:** Suicide attempts and self-directed violence documented in electronic health records by a VHA provider. Hazard ratios (HR) for time to first suicide attempt and self-directed violence (separate models) for the infected versus comparator group were measured using Cox regression models. Analyses were performed for short-term (days 1-30), long-term (days 31-365) and one-year (days 1-365) and further stratified by age and prior self-directed-violence history. Sensitivity analyses included censoring to address comparators crossing over by later testing positive for SARS-CoV-2.

**Results:** Among the 1,190,974 Veterans included, during the one-year period after the index date; 3,078 (0.258%) had a suicide attempt and 2,887 (0.242%) had self-directed violence. Regardless of follow-up duration, the HRs for suicide attempts and self-directed violence were higher for the infected group. For suicide attempts, short-term HR=2.54 (95% Confidence Interval [CI]: 2.05 to 3.15), long-term HR=1.30 (CI: 1.19 to 1.43) and one-year HR= 1.41 (CI: 1.30, 1.54). For self-directed violence, short-term HR=1.94 (CI: 1.51 to 2.49), long-term HR=1.32 (CI: 1.20 to 1.45), and one-year HR=1.38 (CI:1.26, 1.51).

**Conclusions and Relevance:** In matched cohorts, Veterans who had a positive SARS-CoV-2 PCR test had a higher risk of suicide attempt and self-directed violence that were greatest within the first 30 days and present for at least one year following. These findings highlight the importance of assessing patient experiences of suicide attempt and other forms of self-directed violence during different time periods post-infection to identify opportunities to augment prevention efforts and support those affected.

**Trial Registration:** Not applicable

**Key Points:** *Question:* What were the risks of suicide attempts and self-directed violence among US Veterans with SARS-CoV-2 infection compared to a matched cohort?

*Findings:* In this target trial emulation study of a nationwide observational cohort of 1,190,974 matched US Veterans in the Veterans Health Administration from 3/1/2020-3/31/2021, those with a confirmed PCR test for SARS-CoV-2 infection had increased risk of both suicide attempts and self-directed violence that was greatest within 30 days after infection and persisted over the following year. Over the year, those in the infected group had 1.40 times risk of a suicide attempt and 1.38 times risk of experiencing self-directed violence versus those in the comparison group.

*Meaning:* COVID-19 survivors may require additional screening and prevention resources for suicide attempts and other forms of self-directed violence.

## Introduction

Over 100 million people in the United States have been infected with SARS-CoV-2 and developed COVID-19 illness.^1^ There have been mixed findings on studies of rates of suicide-related outcomes during the pandemic,^2–4^ but it is clear that the effects of both SARS-CoV-2 infection and the pandemic itself contribute to poor mental health outcomes more generally. Distinguishing between these two potential causes—the effects of the pandemic and the effects of infection with SARS-COV-2—could help to inform suicide prevention efforts.

The US Veteran population—with a high prevalence of suicide risk factors compared with the US population,^5–8^ may be especially at risk for self-directed violence due to SARS-CoV-2 infection.^3,9,10^ Suicide prevention is the top clinical priority of the US Veterans Health Administration (VHA), which serves 9+ million Veterans each year.^11^ Since suicide attempts are one of the strongest known predictors of future suicide—up to a decade later—identifying veterans who have attempted suicide or engaged in self-directed violence is important in the context of the COVID-19 pandemic.^8,12,13^ Although suicide prevention efforts have been intensified during the pandemic,^14,15^ the association between SARS-CoV-2 infection and suicide attempts or self-directed violence, remains poorly understood.

To address this gap, we utilized data from the VHA COVID-19 Observational Research Collaborative (CORC), which has assembled extensive electronic health record (EHR) datasets of SARS-CoV-2 infected and matched uninfected comparator cohorts.^16–18^ We used an emulated trial design (eTable 1) to evaluate the association of SARS-CoV-2 infection on suicide attempts and other forms of self-directed violence among VHA patients.^17–20^

## Methods

### Study population

We included Veterans in VHA care who had at least one VHA-based positive SARS-CoV-2 PCR test (infected) and a matched group of similar patients without a documented SARS-CoV-2 infection (comparator) on or before their matched infected patient’s date of infection (index date). Per the CORC protocol,^17^ we generated matched cohorts utilizing a combination of exact and time-varying propensity score matching based on EHR-derived covariates. Briefly, CORC identified Veterans were receiving care in the VHA system who were and were not infected with SARS-CoV-2 on a rolling monthly basis from March 1, 2020 through April 30, 2021.^17,21^ From an initial 126,689,864 person-months of observation, CORC generated final matched cohorts of 208,536 Veterans infected and 3,014,091 uninfected (comprising 5,173,400 total person-months of follow-up and up to 25 matches) Veterans. There were 5 exact-matched covariates (index month, sex, any immunosuppressive medication use, state of residence, and COVID-19 vaccination status) and 38 propensity-matched covariates (e.g., demographic characteristics, immunosuppressive medications (binary), nursing home residence at any time, diagnosed CDC high-risk conditions (11), body mass index, etc.). Exclusions were as follows: no VHA utilization in prior two years; implausible age, weight, or height; deceased by baseline date, living outside of the 50 states or Washington, D.C.; prior SARS-CoV-2 diagnosis per Medicare claims (for comparators); or lack of a suitable match. Matching was performed by the PSMATCH procedure from SAS/STAT 15.1 in SAS® 9.4M6.^17^ We used the best 5 matches. To ensure a full year follow-up, we included matched infected and comparators through March 31, 2021. Our cohort included 199,474 in the SARS-CoV-2-infected group and 992,036 in the comparator group (Figure 1).

**Figure 1.** STROBE Diagram of Cohort Derivation. **Figure 1 Footnotes:** ^a^ There were 1,222,272 comparators matched to more than one case. In the numbers presented in this figure, comparators that were matched to more than one infected case have been counted as many times as they appeared in the dataset. The number of unique comparators at each step in the flow diagram are presented in the footnotes below. ^b^ Those excluded in the prior matching included: No CAN Score; no primary care, missing height, weight. or implausible value; age missing or implausible; ZIP code missing or not in DC or 50 states; had a Medicare COVID-19 diagnosis; no suitable match. Additionally excluded among the comparators, those that became a “case” in the same month.^17^ ^c^ Unique number of uninfected comparators matched to an infected case N=2,931,099 ^d^ Unique number of matched comparators with unique identifiers: N=2,928,958 ^e^ Unique number of alive matched comparators at index date: N=2,919,231 ^f^ Unique number of matched comparators that were dropped: 2,049,791 ^g^ Unique number of matched comparators used for analysis: N=869,440. Of the comparators included in the analysis, 104,497 were matched to more than one infected case.

### Data Sources

The VHA employs a comprehensive EHR that centralizes data from all VHA facilities, the Corporate Data Warehouse (CDW), including information about VHA purchased community care.^22^ We also used the COVID-19 Shared Data Resource (CSDR),^23^ a set of analytic variables and datasets to facilitate COVID-19 research and operations. COVID-19 vaccination, date of death, and demographic information from VHA-linked Medicare files^24^ were included.

The study was approved by Institutional Review Boards at VHA facilities in Seattle, Portland, Durham, Ann Arbor, and Palo Alto, which waived the requirement to obtain informed consent because this was a retrospective study of existing data.

### Outcome Measures: Suicide Attempts and Self-Directed Violence

Suicide attempts and other forms of self-directed violence are tracked and reported by VHA providers as part of routine clinical care via completion of a Suicide Behavior and Overdose Report (SBOR) or a VHA Comprehensive Suicide Risk Evaluation (CSRE). Both historical and recent events are reported, including some limited details, such as whether the patient was hospitalized for the event. Based on these reports, the VHA Program Evaluation and Resource Center (PERC) creates curated self-directed violence event datasets, including resolving any duplicate reporting, in accordance with VHA Office of Mental Health and Suicide Prevention (OMHSP) event definitions and classifications and using CDC guidelines.^25–28^ A key strength of these PERC-curated data over relying on ICD-10 codes is a more precise event date reconciled through this process, reducing risk of ascertainment bias (i.e., COVID-19 resulting in more visits and more reporting of historical events).

We used the PERC data to identify occurrences of suicide attempts and self-directed violence. We did not include suicide deaths as an outcome because a lag in the availability of cause of death data from the VHA Mortality Data Repository, the gold standard for verifying deaths, meant that these could not be validated.^13,29^ We instead report the frequency of unvalidated suicide deaths as reported within the PERC data. Self-directed violence included preparatory behaviors, non-suicidal self-directed violence, and undetermined self-directed violence. When more than one event type was reported for the same date (n=203 pre- or post-index events), we selected the most severe event type (e.g., suicide attempts selected over preparatory behaviors). If the event date was within two days of the death date, we reclassified as “suicide death” (n=3). We assessed outcomes separately for three periods following the index date: short-term (day 1-30), long-term (day 31-365) and one-year (day 1-365).

### Statistical Analyses

We compared demographic and clinical characteristics between the infected and comparator groups using descriptive statistics (frequencies and percentages for categorical variables, means and standard deviations for continuous variables) and standardized mean differences (SMD).^30,31^ We separately described Veterans with unverified suicide deaths (frequencies only). We calculated events per 100,000 person years (event rates) by dividing the sum of events in the time period by the sum of follow up times (in years, from index date to death, event, or 1 year) and multiplying by 100,000. Event-free follow-up was also assessed using standard Kaplan-Meier plots for each outcome/ and across pre-defined sub-groups.

As an emulated trial design, we used an intent to treat (ITT) approach for our regression analyses. All analyses described below were repeated under a per protocol framework that censored either patients in the comparator group who later had a positive SARS-CoV-2 PCR (PP1) or censored the entire matched group when any member of the comparator group had a positive PCR (PP2).

Our primary analyses used Cox regression models with time to first suicide attempt and time to self-directed violence event as the outcomes and being infected with SARS-CoV-2 or not at the time of the index date as the independent variable. Models censored for death used stratified baseline hazard specifications, with the matched groups as the strata and robust error estimation clustered on the matched groups. No adjustment variables were included since the SMD’s were less than 0.1 for all matched variables (See Table 1), following evidence from Nguyen et al^32^ there is negligible additional benefit of covariate adjustment with minimal imbalance after matching.^32,33^ Stratified analyses were performed to estimate hazard ratios (HR) for specific age groups (18-45, 46-59, 60-64, 65-74, and 75 years or older). Since risk of future suicide events is higher in patients with a history of prior suicide attempts or self-directed violence,^13^ we also stratified analyses by suicide attempt or self-directed violence two years pre-index.

**Table 1.**
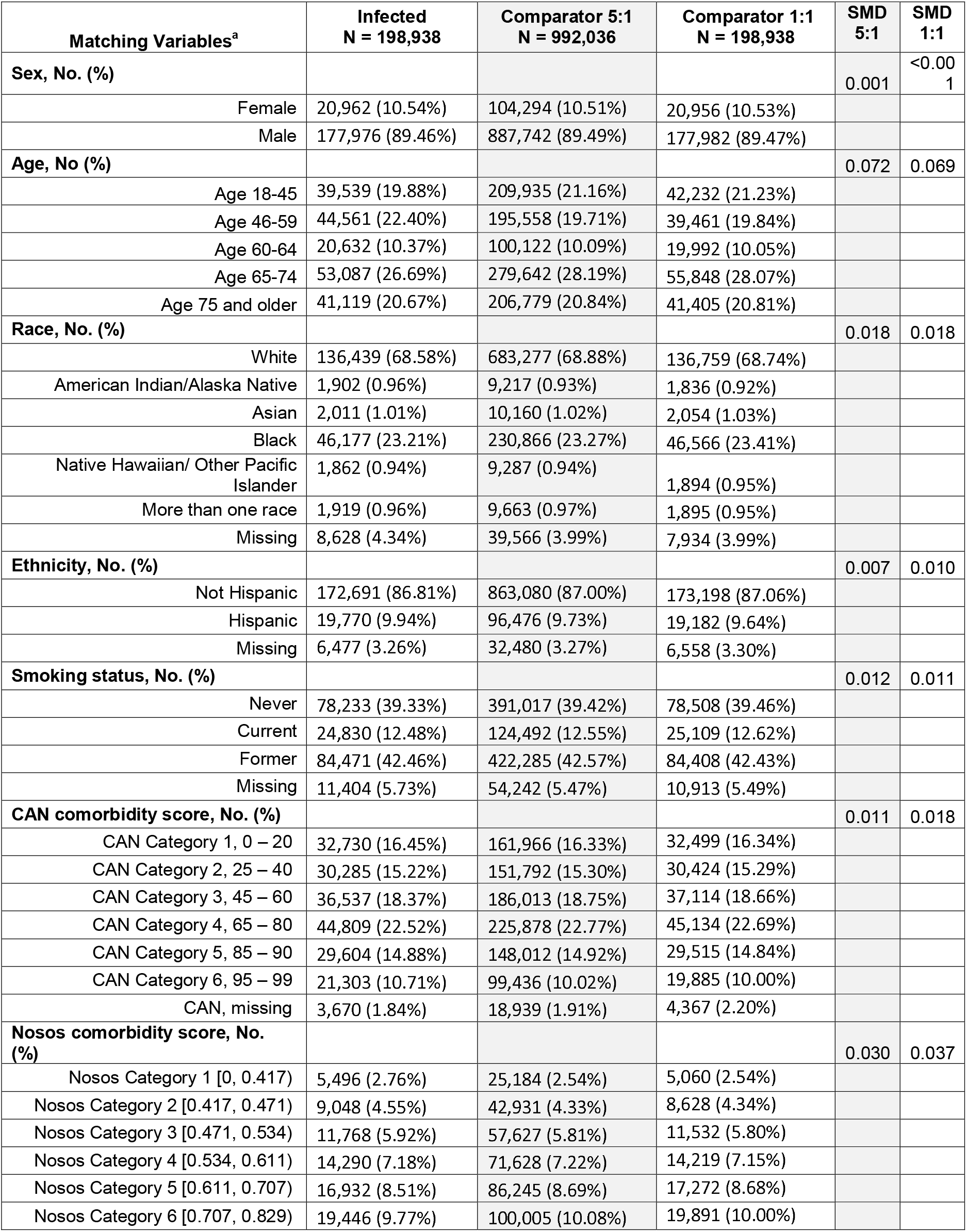
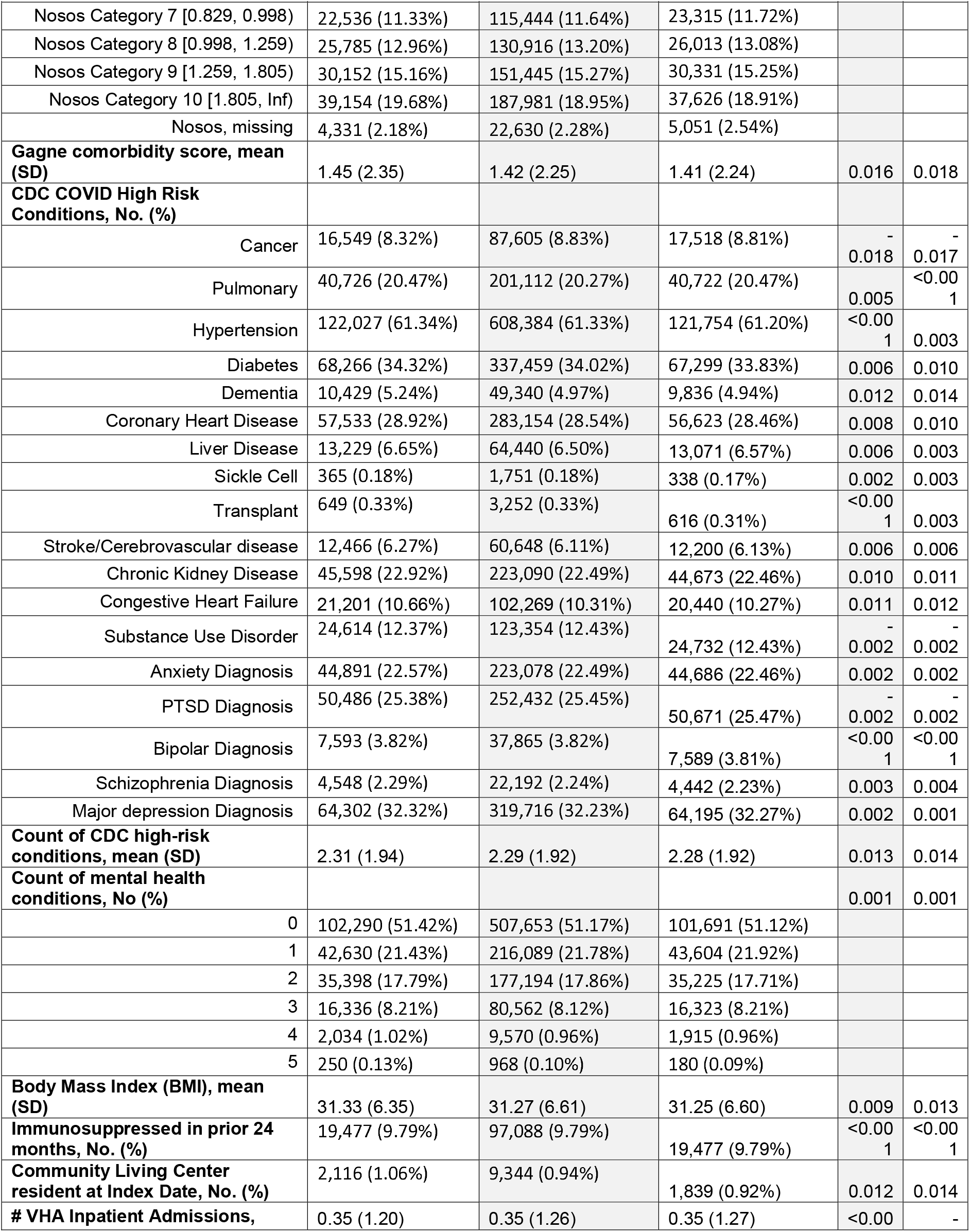
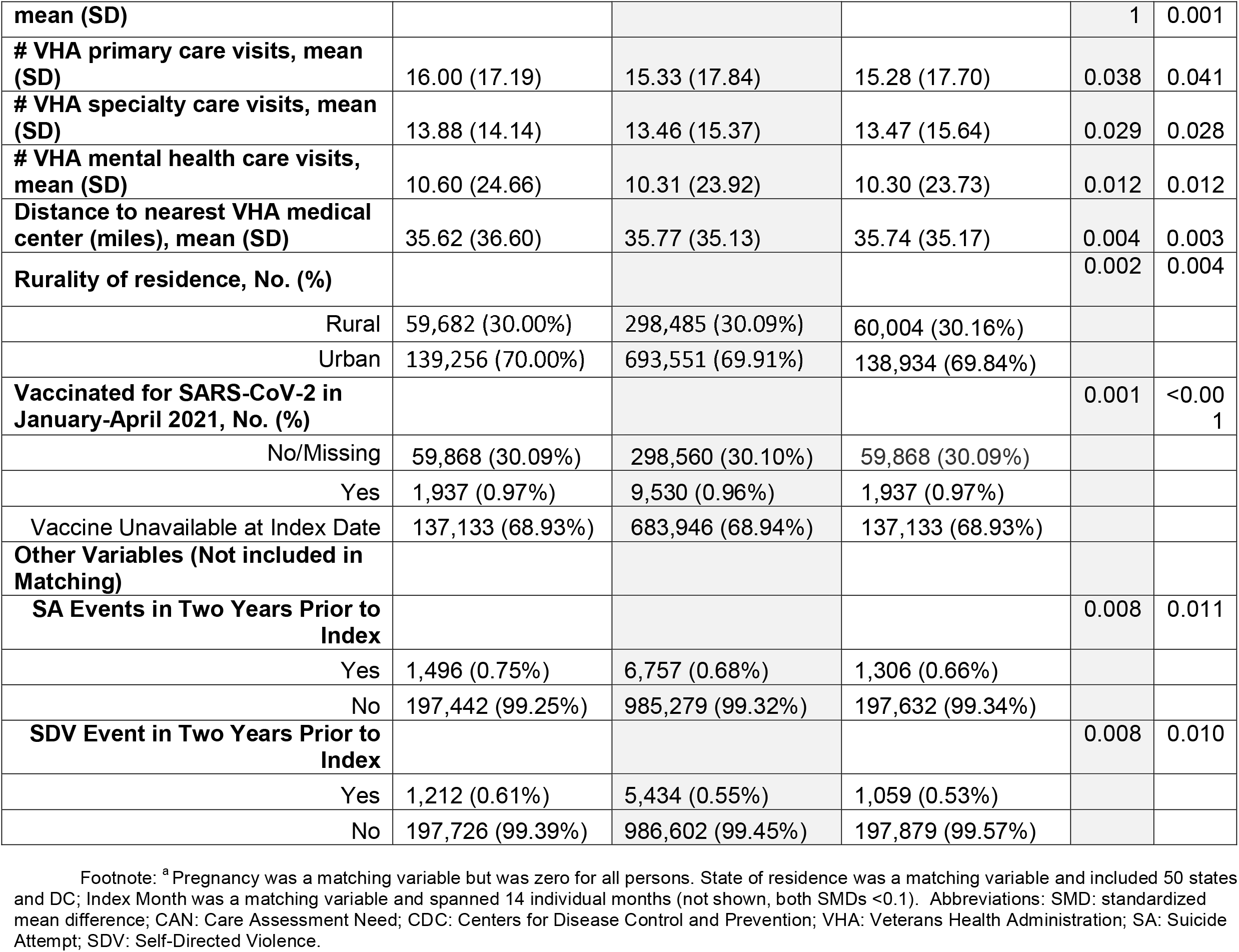
Baseline Characteristics of Veterans in the Infected and Matched Comparator Groups for the 5:1 and 1:1 Matched Cohorts, March 1, 2020 – March 31, 2021.

In sensitivity analyses, we used generalized estimating equations (GEE) with the 1:1 matched comparators to compare the odds of having at least one suicide attempt (separately for self-directed violence) during each of the follow-up periods,^34,35^ with clustering by matched pair and an independent covariance structure. As before, models included no other adjustment variables, and the same stratified analyses were performed. For GEEs we applied the same two per protocol analyses PP1 and PP2, as well as excluding deaths in either group (PP3) and excluding entire pairs with crossover or death (PP4).

All regression analyses were conducted with R [R version 4.1.2, the survival R package version 3.4.0 for Cox regression models and the geepack R package version 1.3.9 for GEE models.]

## Results

Among the 1,190,974 Veterans in our overall matched cohort, there were 198,938 Veterans in the infected group and 992,036 Veterans in the comparator group. Demographic and other characteristics of the Veterans in the infected and matched comparator groups were similar (SMD < 0.1) (Table 1).

Within one year of the index date, 3,078 (0.258%) cohort members had at least one documented suicide attempt and 2,887 (0.242%) at least one documented self-directed violence (Table 2), an incidence of 267 and 250 events per 100,000 person-years, respectively. The unadjusted event rates for both suicide attempts and self-directed violence were highest in the first 30 days, but event rates were higher in the infected group than among comparators for all follow-up durations (Table 2).

**Table 2.** Description of Suicide attempts and Self-Directed Violence Among All Infected and Matched Comparators for Short Term, Long Term, and One-Year Follow-up Periods After Index Date, for the 5:1 Match, through March 31, 2022.

Footnote: Abbreviations: PY: person-years
|  | Short-Term Outcomes<br>(Day 1-30 Post Index) |  |  |  | Long-Term Outcomes<br>(Day 31-365 Post Index) |  |  |  | One-Year Outcomes<br>(Day 1-365 Post-Index) |  |  |  |
| --- | --- | --- | --- | --- | --- | --- | --- | --- | --- | --- | --- | --- |
|  | Overall | Infected | Comparators | Percent<br>Difference | Overall | Infected | Comparators | Percent<br>Difference | Overall | Infected | Comparators | Percent<br>Difference |
|  | N =<br>1,190,974 | N =<br>198,938 | N =<br>992,036 |  | N =<br>1,190,974 | N =<br>198,938 | N =<br>992,036 |  | N =<br>1,190,974 | N =<br>198,938 | N =<br>992,036 |  |
| **Suicide Attempt, No. (%)** | 382<br>(0.03%) | 127<br>(0.06%) | 255<br>(0.03%) | 0.03% | 2,778<br>(0.23%) | 555<br>(0.28%) | 2,223<br>(0.22%) | 0.06% | 3,078<br>(0.26%) | 657<br>(0.33%) | 2,421<br>(0.24%) | 0.09% |
| **Events/**<br>**100,000 PY** | 393 | 798 | 313 |  | 263 | 328 | 250 |  | 267 | 355 | 250 |  |
| **Self-Directed Violence, No. (%)** | 309<br>(0.03%) | 85<br>(0.04%) | 224<br>(0.02%) | 0.02% | 2,636<br>(0.22%) | 534<br>(0.27%) | 2,102<br>(0.21%) | 0.06% | 2,887<br>(0.24%) | 606<br>(0.31%) | 2,281<br>(0.23%) | 0.08% |
| **Events/**<br>**100,000 PY** | 318 | 534 | 275 |  | 249 | 315 | 237 |  | 250 | 327 | 235 |  |

Kaplan-Meier plots showing the event-free follow-up probability for the one-year period for suicide attempt and self-directed violence and for the corresponding age stratifications are shown in Figure 2.

**Figure 2.** Intent to Treat Cox Regression Model for Suicide Attempts & Self-Directed Violence for Short-Term (Day 1-30), Long-Term (Day 31-365) and One Year (Day 1-365) Follow-up Periods After Index for the 5:1 Matched Cohort. † 95% Confidence intervals >5 are not shown for clarity of overall figure or would not converge where line is missing. Upper CI limits are as follows: for Suicide Attempts: Short-Term, Age 60-64: 11.64; Short-Term, Age 65-74: 11.94; Short-Term, Age 75+: 97.26; Short-Term, Previous SA: 22.06; Previous SDV: 22.06, 12.07, 7.26, Short-Term, Long-Term, Combined, respectively. For Self-Directed Violence: Short-Term, Age 60-64: 19.38; Short-Term, Age 65-74: 7.39; Short-Term, Age 75+: 13.69; Previous SA: Long-Term and Combined: 5.61 and 5.61, respectively; Previous SDV: Long-Term and Combined: 6.26 and 6.26, respectively. Models for Self-Directed Violence (Previous SA/SDV) would not converge. For the Short-Term, Age 75+ Suicide Attempt outcome, the HR was omitted for figure clarity: HR 10.22. Total N=1,119,974; Age 18-45 N=249,474; Age 46-59 N=240,119; Age 60-64 N=120,754; Age 65-74 N=332,729; Age 75+ N= 247,898; Previous SA N=8,253; No Previous SA N=1,182,721; Previous SDV N=6,646; No Previous SDV N=1,184,328.

There were 237 suicides reported in PERC data among cohort members during the observation period, with no significant differences in the one-year incidence of suicide deaths between the infected and comparator groups: 41 (0.021%) vs. 196 (0.020%), p=0.81 respectively. Most suicides occurred more than 30 days after the index date (n=175) rather than within 30 days (n=32), although these data are exploratory since cause of death could not be verified.

### Cox Regression Analyses

#### Suicide Attempts

In the ITT Cox regression analyses, the risk of a suicide attempt in the infected group versus the comparators was higher in all follow-up periods: short-term Hazard Ratio (HR)=2.54; (95% Confidence Interval [CI] 2.05-3.15; P<0.001), long-term HR=1.30; (CI 1.19-1.43; P<0.001) and one-year HR=1.41; (CI 1.30-1.54; P<0.001). (Figure 2, eTable 3).

In age group-stratified ITT analyses (ages 18-45, 46-59, and 65-74 years), the risk of suicide attempt was higher among those in the infected versus the comparator group during all three time-periods, with the highest rates in the short term. For two age groups (60-64; and 75 and older years) the risk of suicide attempt was significantly higher for the infected group only in the short term.

In ITT analyses stratified by prior suicide attempt or self-directed violence, the risk of suicide attempt for the infected group was higher for those with no prior suicide attempt or other self-directed violence in all three periods. Among those with prior self-directed violence there was no significant difference for the infected versus comparators, although sample sizes were small in these subgroups.

#### Self-Directed Violence

In the ITT Cox regression analyses the risk of self-directed violence in the infected group versus the comparators was higher in all follow-up periods: short-term HR=1.94 (CI 1.51-2.49; P<0.001); long-term HR=1.32 (CI 1.20-1.45; P<0.001); and, one-year HR=1.38 (CI 1.26-1.51; P<0.001). (Figure 2; eTable 3).

In age-stratified ITT analyses amongst 18–45-year-olds, the risk of self-directed violence was higher for the infected group in all three time periods, while those 65-74 years and 75 years and older the infected groups had higher self-directed violence risk only for the long-term and one-year follow-up periods.

In ITT analyses stratified by prior self-directed violence, the risk of self-directed violence for the infected group was higher for those with no prior suicide attempt or other self-directed violence in all three periods. Among those with prior suicide attempt or other self-directed violence, there was no significant difference for the infected versus comparators, although sample sizes were small in these subgroups and the short-term models did not converge. (eTable 3)

The results of per protocol sensitivity and secondary GEE analyses were largely similar to the primary analyses. (eTable 4-6)

## Discussion

Our findings demonstrate that those infected with SARS-CoV-2 were at increased risk for attempted suicide and self-directed violence compared with matched controls. For up to one-year post-exposure, suicide attempts and self-directed violence among Veterans in the VHA were higher for those in the infected group compared to similar matched Veterans without infection. This investigation is important as a national US study reporting on the incidence of suicide attempts and self-directed violence among Veterans, a population with elevated risk of suicide, with contemporaneous, well-matched controls. Contemporaneous controls are important to examine the impact of infection on mental health outcomes, given the multiple other stressors co-occurring with infection in the pandemic. While research continues about the underlying neuropsychological and other mechanisms of SARS-CoV-2 infection and its sequelae,^36^ greater understanding of the patient experience of exposure is also vital.

We found that those in the infected group had 1.41 times higher risk of a suicide attempt and 1.38 times higher risk of self-directed violence versus those in the comparator group within one year after the index infection date. In comparison, the National Health and Resilience in Veterans Study, which followed a cohort of 3,078 Veterans from November 2019 to December 2020, reported that Veterans with self-reported COVID-19 were 2.41 times more likely than uninfected participants to report suicidal ideation.^3^ Our study relied on PCR confirmed SARS-CoV-2 infection, distinguished historic from recent self-directed violence, and includes a longer time period than this earlier work. Our results support the need for continued monitoring to facilitate recognition of suicide attempts and self-directed violence and referral for appropriate care.^37^

Among three of our age groups (ages 18-45, 46-59, and 65-74 years) suicide attempts were higher among those infected with SARS-CoV-2 than among comparators. A meta-analysis of 13 studies that assessed suicide attempts conducted early in the pandemic reported that the prevalence of suicide attempts decreased as age increased, although most were cross sectional and mainly involved younger age groups.^38^ Our results over a longer period suggest both younger and older adults experience suicide attempts and self-directed violence.

Studies of temporal trends in suicide risk during the course of infectious pandemics are lacking.^39^ We found that HR for suicide attempts and self-directed violence were greatest during the first 30 days following the infection date. This was also true in many subgroups. For example, among those aged 65-74, for the risk within 30 days for suicide attempts for those infected were 4.10 times that of comparators, whereas the risk from day 31-365 was 1.58 times that among comparators. Some suggest bolstering social connectedness and protective psychosocial characteristics may help mitigate risk,^40,41^ and the timing of such actions may be important. Our results call for more detailed context of suicide attempts and self-directed violence occurring in the immediate aftermath of SARS-CoV-2 infection to identify credible opportunities for prevention.

Prior research has shown that VHA patients with a prior suicide attempt are at increased risk for suicide for more than a decade following the initial attempt.^13^ We thus expected to find risk concentrated among those with a pre-index risk of suicide attempts or self-directed violence. While absolute rates of suicide attempts and self-directed violence were highest among those with a prior history of these events, the relative hazard of suicide attempt or self-directed violence was actually highest among those without prior history of such events.

However, analyses were underpowered for the group with prior experiences, and we cannot conclude there was not an effect.

Overall, although suicide attempts and self-directed violence are infrequent, they are risk factors for future self-directed violence, suicide attempts and suicides.^13,42^ Among Veterans in our cohort, 3,078 (0.258%) had at least one suicide attempt and 2,887 (0.242%) experienced at least one episode of self-directed violence in the year following the index date. It is noteworthy that these unadjusted rates are lower compared to the prior two years (0.693% and 0.558%, respectively for suicide attempt and self-directed violence) among cohort members. It is possible that the overall lower reported rates of suicide attempts and self-directed violence during the pandemic was the result of care disruption and limited access to routine health visits that provide an opportunity to uncover these events.^43–46^ The rates of suicide attempts in a regional study among Veterans at eight VHA facilities in the Southwestern US^47^ that examined trends during the first year of the pandemic (March 13, 2020-March 12, 2021) compared to the prior year produced results similar to ours by showing a decline in suicide attempts, but only overall results were reported without stratification by whether COVID-19 occurred. Collectively, our results highlight the importance of suicide prevention efforts during the pandemic, particularly among those infected with SARS-CoV-2 and beginning early after infection.

There are limitations to our study. First, although we used a dataset that differentiated between current and historical events, we cannot rule out ascertainment bias due to the possibility that patients infected with SARS-CoV-2 infection may have had increased access to healthcare during the pandemic. Second, although we included both VHA and Medicare claims data to identify those who had a PCR-confirmed SARS-CoV-2 infection, it is also possible that some Veterans who may have later become infected, had no health system contact. This potential bias may be accentuated with greater use of home antigen testing, or perhaps situations where patients did not know they were infected.^48^ The waning of the association over time could reflect undetected crossovers from the comparator group. Third, although we achieved close matching on available covariates, our results may still be subject to unobserved confounding.

## Conclusions

This study is the first to our knowledge to employ an emulated trial design to assess the risk of attempted suicide and self-directed violence among patients infected with SARS-CoV-2 compared to a contemporaneous matched uninfected comparator group. Our findings demonstrate that Veterans infected with SARS-CoV-2 were at greater risk for suicide attempts and self-directed violence compared to similar patients who were not infected. Our results indicate that risk for self-directed violence was especially high during the first 30 days after infection but persisted for at least a year after infection. Our findings highlight the importance of assessing the experiences of patients with suicide attempts and self-directed violence beginning early after COVID-19 illness to identify opportunities to augment prevention efforts and improve support for those affected.

## Data Availability

The datasets generated and/or analyzed during the current study are not publicly available due to Department of Veterans Affairs data restrictions prohibiting sharing. Contact the corresponding author, Dr. Denise Hynes, for data requests.

## Funding Acknowledgements

This work was supported by funding from the US Department of Veterans Affairs (VA) Health Services Research and Development (HSR&D) Service awards for the COVID Observational Research Collaborative (C19-21-278; C19-21-279); and for use of the VA-linked Medicare data (SDR 02-237; SDR 98-004).

Drs. Hynes and Maciejewski were supported in part by a VA Research Career Scientist Award (RCS 10-391 and RCS 21-136, respectively).

Dr. Chen was supported by a VA Career Development Award.

The authors all have employment or compensation arrangements with the US Department of Veterans Affairs.

All statements in this article, including its findings and conclusions, are solely those of the authors and do not necessarily represent the views of the US Department of Veterans Affairs or the US government, Oregon State University, Oregon Health & Science University, Portland State University, Duke University, the University of Washington, Johns Hopkins University, and the University of Michigan. No copyrighted material was included in this article.

We also appreciate the data made available by the VHA Office of Mental Health and Suicide Prevention/Program Evaluation and Resource Center.

